# Ontology-based expansion of virtual gene panels to improve diagnostic efficiency for rare genetic diseases

**DOI:** 10.1101/2024.10.31.24315563

**Authors:** Jaemoon Shin, Toyofumi Fujiwara, Hirotomo Saitsu, Atsuko Yamaguchi

## Abstract

**Background:** Virtual Gene Panels (VGP) comprising disease-associated causal genes are utilized in the diagnosis of rare genetic diseases to evaluate candidate genes identified by whole-genome and whole-exome sequencing. VGPs generated by the PanelApp software were utilized in a UK 100,000 Genome Project pilot study to filter candidate genes, thus enhancing diagnostic efficiency for rare diseases. However, PanelApp also filtered out disease-causing genes in nearly 50% of the cases.

**Methods:** Here, we propose various methods for optimized approach to design VGPs that significantly improve the diagnostic efficiency by leveraging the hierarchical structure of the Mondo disease ontology, without excluding disease-causing genes. We also performed computational experiments on an evaluation dataset comprising 74 patients to determine the optimal VGP design method.

**Results:** Our results demonstrate that the proposed method can significantly enhance rare disease diagnosis efficiency by automatically identifying candidate genes. The proposed method successfully designed VGPs that improve diagnosis efficiency without excluding disease-causing genes.

**Conclusion:** We have developed novel methods for VGP design that leverage the hierarchical structure of the Mondo disease ontology to improve rare genetic disease diagnosis efficiency. This approach identifies candidate genes without excluding disease-causing genes, and thereby improves diagnostic efficiency.

## Background

Rare diseases exhibit an extremely low prevalence rate, estimated to be around 10,000 cases worldwide [1]. The estimated number of individuals affected by these diseases worldwide is estimated to surpass 400 million, with a significant portion remaining undiagnosed for years [2]. Approximately 80% of the rare diseases are believed to have a genetic origin [1]. Next-generation sequencing (NGS) have reduced the cost and time required to decode genetic sequences, and thus provides with a powerful tool for diagnosing rare diseases as well [3]. However, diagnosing rare diseases using NGS still requires a labor-intensive process of literature search to identify a single candidate disease-causing gene associated with disease symptoms. Even for experts, this manual interpretation may take hours [4].

Virtual gene panels (VGP) comprising a set of disease-related causal genes are used to identify candidate genes for disease diagnosis efficiency and thus to facilitate the interpretation process. The number of potential candidates can be significantly reduced by considering the overlap between candidate genes and the set of genes in the VGP corresponding to an initial diagnosis. To this end, a pilot study of UK 100,000 Genomes Project for rare disease diagnosis developed PanelApp, a resource comprising 340 VGPs that can be applied for the manual interpretation of whole-genome sequencing results [5]. The PanelApp is a publicly available knowledge base that allows virtual gene panels to be created, stored, and queried. It also includes a crowdsourcing tool that allows genes, short tandem repeats, and copy number variants to be added or reviewed by experts in the international scientific community. Manual curation in this way provides an opportunity to establish standard virtual gene panels with sufficient evidence on disease association [6].

PanelApp was efficient in identifying candidate genes in half of the cases, yet also filtered out disease-causing genes in the remaining half [7]. This may be attributed to two factors. First, PanelApp includes only 340 VGPs that are identified via manual curation. This limited set of genes may not necessarily be relevant for an initial diagnosis. Second, if the initial diagnosis is slightly incorrect, the corresponding VGP may not include the disease-causing gene in the respective case.

In this study, we propose a method for automatically selecting VGPs developed independently using a knowledge graph. Additionally, we propose several methods to expand VGPs using the hierarchical structure of the Mondo disease ontology (Mondo) [8] in cases where the initial diagnosis is slightly incorrect. The basic premise of the expanded VGP design is the addition of genes associated with the superclass or siblings of the disease initially diagnosed in Mondo to an original set of genes. We evaluated the diagnostic efficiency of these methods using 74 cases of rare genetic diseases. Our results showed that the best method successfully identified candidate genes while retaining disease-causing genes with high probabilities.

## Methods Knowledge graph

We collected a large amount of medical data and integrated it into the RDF Portal (https://rdfportal.org/) [9]. Then, we constructed a resource description framework (RDF) based knowledge graph with interoperability based on this data. The knowledge graph included more than 16 million triples with 15 properties. Using this knowledge graph, we constructed a process for the design of VGPs (https://integbio.jp/rdf/dataset/pubcasefinder). Fig. 1 illustrates a simplified knowledge graph derived from the original, with a focus on highlighting the paths connecting diseases to genes.

**Fig. 1.**
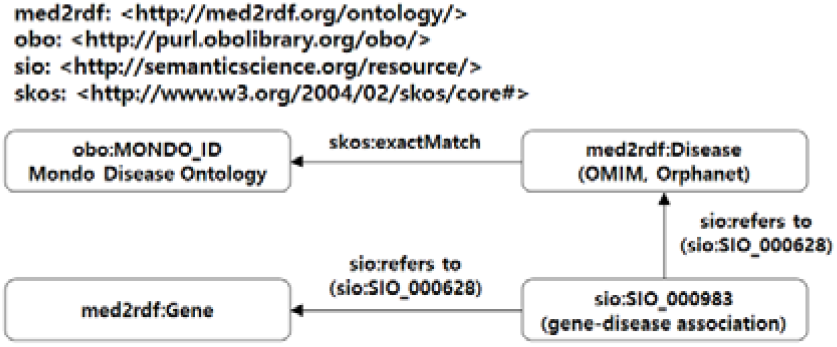
The knowledge graph for designing VGPs automatically

All diseases and genes were defined as instances of “med2rdf:Disease” and “med2rdf:Gene” classes, respectively, by Med2RDF-ontology (http://med2rdf.org/) [10]. Gene-disease associations were defined as instances of the “sio:SIO_000983” class as defined by Semantics Science Integrated Ontology [11].

We collected gene-disease associations from the following sources.

1. MIM2Gene (https://ftp.ncbi.nlm.nih.gov/gene/DATA/mim2gene_medgen). Associations between MIM Numbers with type “phenotype” and NCBI Gene IDs were extracted.
2. OrphaData (http://www.orphadata.org/data/xml/en_product6.xml). Associations between OrphaCodes and Gene symbols were extracted.
3. GenCC (https://search.thegencc.org/download) [14]. Associations between disease IDs, including MIM Numbers, OrphaCodes, and Mondo IDs and HGNC IDs, were extracted.

We matched MIM numbers in Online Mendelian Inheritance in Man [12] and OrphaCodes in Orphanet Rare Disease Ontology [13] with their corresponding Mondo IDs using the “equivalentTo” classes in Mondo (data-version:releases/2021-06-01). In the constructed knowledge graph, genes were thus associated with a disease ID in Mondo through med2rdf:Disease and sio:SIO_000983. We defined a set of genes associated with a disease using this graph. For disease ID *d* in Mondo, we denote a set of genes by *G*_0_(*d*) connected to *d* using the path in the knowledge graph. If *d* has children, the sets of genes associated with them are also considered. We define *G*(*d*) = *G*_0_(*d*)∪ (∪ *d′* _*is a subClassof d*_*G*(*d*′))as a set of genes associated with disease *d*.

## Methods for expanding VGPs

Initially, when disease *d* is diagnosed, *G*(*d*)can be regarded as a VGP for *d*. **Original set (OR):** For a given disease *d*, output a set *G*(*d*)of genes. Using the OR, we can obtain as many VGPs as the number of Mondo IDs.

However, similarly to the issue mentioned earlier, OR excludes disease-causing genes if the initial diagnosis is slightly different. Here, we present six methods for expanding the VGPs initially included in the OR to overcome this problem.

Mondo forms a directed acyclic graph, with nodes representing diseases and edges “rdfs:subClassOf” relationships between diseases. Using this graph, we expanded the VGPs based on OR by adding superclass genes to Mondo using these methods. Four of these methods involve nodes ascending once or twice through the RDF property:subClassOf. The remaining two methods involve ascending nodes until the number of genes exceeded a certain threshold.

Here, we describe four simple rolling-up methods to expand the VGP using rdfs:subClassOf. **One class up for all paths (1UAP):** For disease *d*, compute a set of genes *G* ={*g*|*g* ∈ G(*d*′), *d rdfs*:*subClassOf d*′}. **Two classes up for all paths (2UAP):** For disease *d*, compute a set of genes *G* ={*g*|*g* ∈ *G*(*d*′′), *d rdfs*:*subClassOf d*′ *and d*′ *rdfs*:*subClassOf d*′′}. **One class up for minimum paths (1UMP):** For disease *d*, compute a set of genes *G* =*argmin*_|*G*(*d*′)|_ {*g*|*g* ∈ *G*(*d*′), *d rdfs*:*subClassOf d*′}. **Two classes up for minimum paths (2UMP):** For disease *d*, compute a set of genes, *G* =*argmin*_|G(*d*′′)|_ {*g*|*g* ∈ *G*(*d*′′), *d rdfs*:*subClassOf d*′ *and d*′ *rdfs*:*subClassOf d*′′}. Let us suppose that the initial disease *d* is schizencephaly based on the example shown in Fig. 2B. In the 1UAP method, we roll up *d* to its one-step higher classes in all paths, resulting in *d* being changed to an encephaloclastic disorder or congenital nervous system disorder. Similarly, in the 2UAP method, we rolling-up *d* to its two-step higher classes, leading to *d* being changed to cerebral malformation and nervous system disorders. Thus, the 1UAP method expands the VGP set for *d* by including genes associated with Schizencephaly, Encephaloclastic disorders, and Congenital nervous disorders. In contrast, the 2UAP method expands the VGP set to include genes associated with Schizencephaly, Encephaloclastic disorders, congenital nervous system disorders, cerebral malformations, and nervous system disorders. With 1UMP, *d* is changed to encephaloclastic disorder, the one with the lowest number of genes among encephaloclastic disorder and congenital nervous system disorder, which is a one-step higher class. With 2UMP, *d* is changed to cerebral malformation, the one with the lowest number of genes among cerebral malformation and nervous system disorder, which is a two-step higher class.

**Fig. 2.**
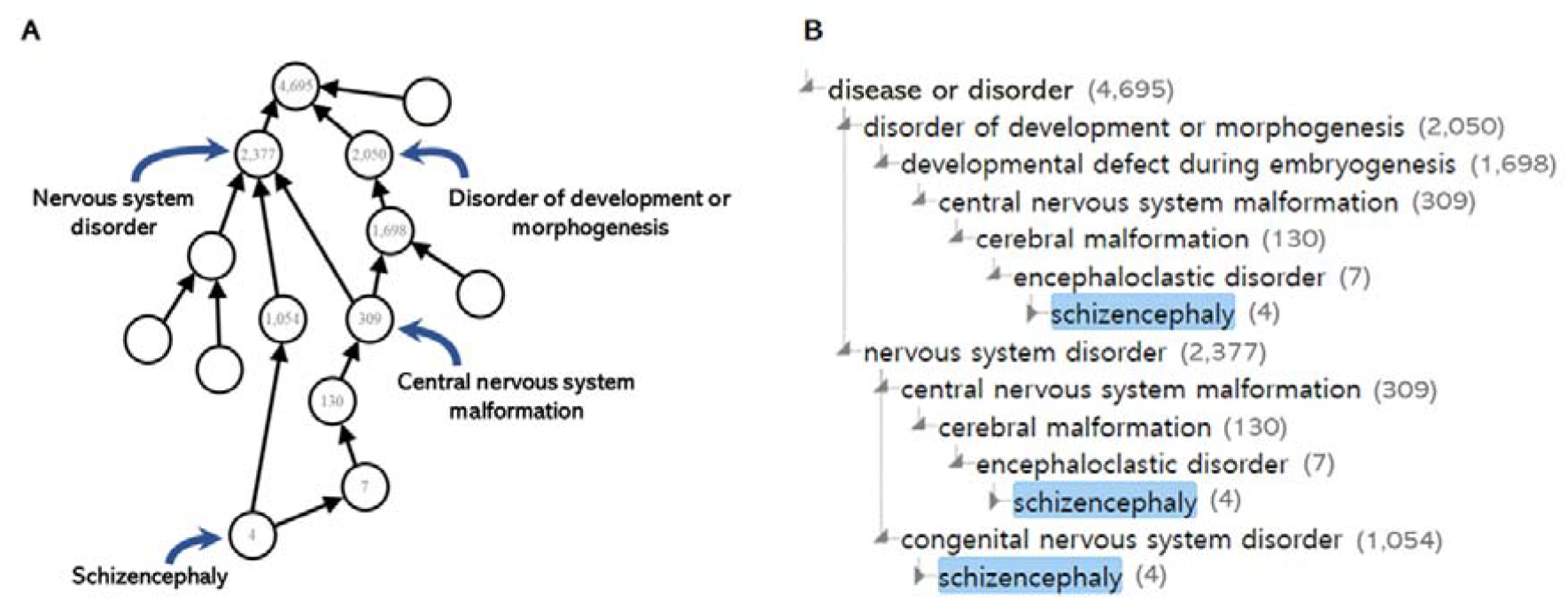
Examples A and B are provided as illustrative examples to aid in understanding the structure of Mondo. An example of Schizencephaly (MONDO_0010011). The numbers next to the Disease in Fig. 2B represent the count of genes that are combined in the union of the gene set of that Disease and the gene set of its subclasses.

Additionally, we describe the following methods to expand the VGP by rolling-up until the number of genes exceeds the threshold *k*.

- All paths up with threshold *k* (TH *k* AP)
- Minimum path up with threshold *k* (TH *k* MP)

For an initially diagnosed disease *d*, TH *k* AP and TH *k* MP compute a set *D* of Mondo IDs, such that each *d*′ in *D* is *d* or an ancestor of *d* and the size of *G*(*d*′)is equal to or greater than the threshold *k*. TH *k* AP outputs the union of *G*(*d*′)for all *d*′ in *D*, and TH *k* MP outputs *G*(*d*′)for *d*′ with a minimum size of *G*(*d*′). The algorithms of TH *k* AP and TH *k* MP are presented in Algorithms 1 and 2, respectively.

### Algorithm 1

TH *k* AP

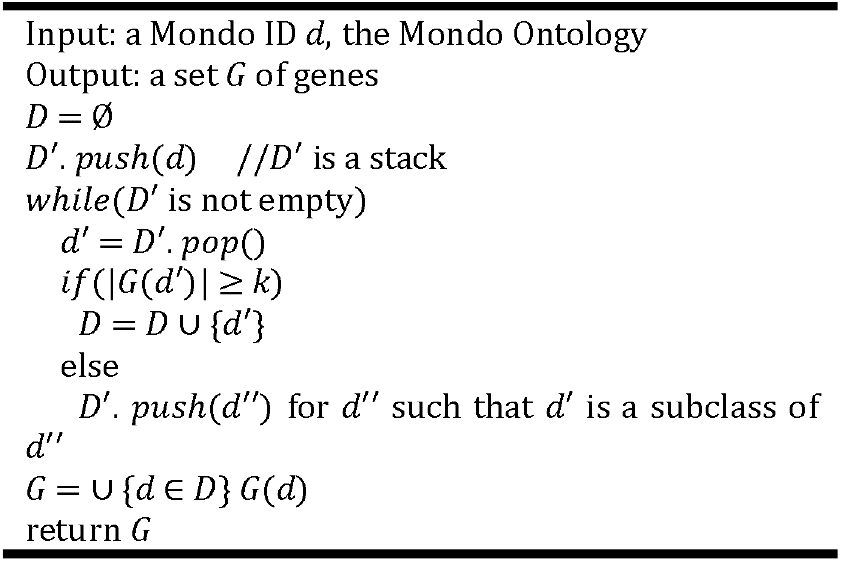

### Algorithm 2

TH *k* MP

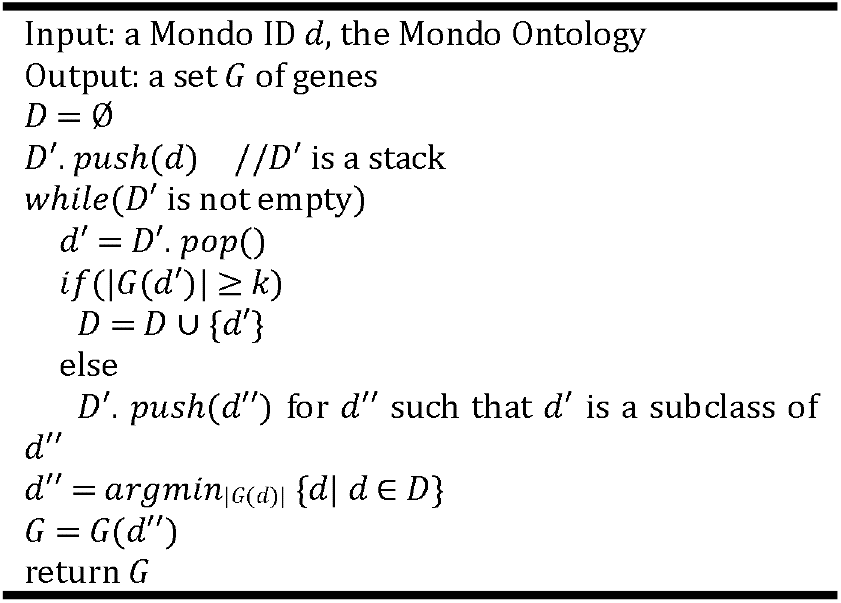

For example, with the initial disease *d* as schizencephaly and the threshold value as 1,000, the TH *k* AP method results in changing *d* to a congenital nervous system disorder and developmental defects during embryogenesis, which have gene counts exceeding the threshold. However, the TH *k* MP method changes *d* change to congenital nervous system disorder, which is the class with the smallest number of associated genes among congenital nervous system disorders and developmental defects during embryogenesis.

## Results

We evaluated the diagnostic efficiency of the developed VGPs using a whole-exome sequencing (WES) dataset comprising 74 patients. The schema of the test dataset consists of “Patient ID”, “Initial diagnosis (using Mondo)”, “Candidate gene list”, “Phenotypes (using HPO)”, and “Final disease-causing genes”. The median and average number of candidate genes were 384 and 383.25, respectively. Only a single disease-causing gene was included per patient in *G*. The number of initially diagnosed diseases ranged between one and three, with a median of one.

Using a knowledge graph, the VGPs for approximately 10,000 diseases included in Mondo can be automatically designed based on the OR method. To evaluate the performances of the expansion methods, we computed their coverage and median. Additionally, we evaluated the expectations, considering how extensive the analysis should be when using the panel in practice to determine the final disease-causing genes. The coverage refers to the ratio of VGPs, including disease-causing genes. The median is the median of the sizes of the intersections between the patient’s candidate genes and the VGPs. Expectation is the expected number of genes to be analyzed to determine the final disease-causing gene. If VGPs include disease-causing genes, only the genes at the intersection of the patient’s candidate genes and the VGPs should be analyzed. However, if the VGPs do not include disease-causing genes, all candidate genes should be analyzed. Hence, the expectation was computed using the following formula:

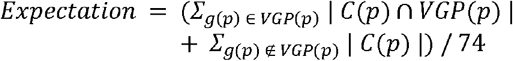

where for patient *p*, a set of genes in the VGP, the disease-causing gene, and a set of candidate genes are denoted as VGP(*p*), *g*(*p*), and *C*(*p*), respectively.

Fig. 3 shows the results for TH *k* AP and TH *k* MP when the threshold was increased from 0 to 4,500 in increments of 250 (All results can be found in Supplementary Material 1). Tables 1 and 2 show the results for TH *k* AP and TH *k* MP when the threshold was increased from 0 to 1,750 in increments of 250.

**Table 1.** The results of the experiments using TH *k* AP.

| Threshold($k$) | 0 | 250 | 500 | 750 | 1,000 | 1,250 | 1,500 | 1,750 |
| --- | --- | --- | --- | --- | --- | --- | --- | --- |
| **Coverage** | 0.6081 | 0.9730 | 1 | 1 | 1 | 1 | 1 | 1 |
| **Median** | 2 | 47 | 60.5 | 62 | 62 | 69 | 69 | 90.5 |
| **Expectation** | 158.15 | 54.93 | 59.19 | 62.04 | 63.73 | 72.80 | 72.80 | 84.20 |

**Table 2.** The results of the experiments using TH *k* MP.

| Threshold($k$) | 0 | 250 | 500 | 750 | 1,000 | 1,250 | 1,500 | 1,750 |
| --- | --- | --- | --- | --- | --- | --- | --- | --- |
| **Coverage** | 0.6081 | 0.9459 | 0.9459 | 0.9730 | 0.9730 | 0.9865 | 0.9865 | 0.9730 |
| **Median** | 2 | 15 | 25 | 29.5 | 30 | 42.5 | 42.5 | 56 |
| **Expectation** | 156.19 | 38.74 | 47.03 | 43.46 | 44.78 | 50.82 | 50.82 | 66.86 |

**Fig. 3.**
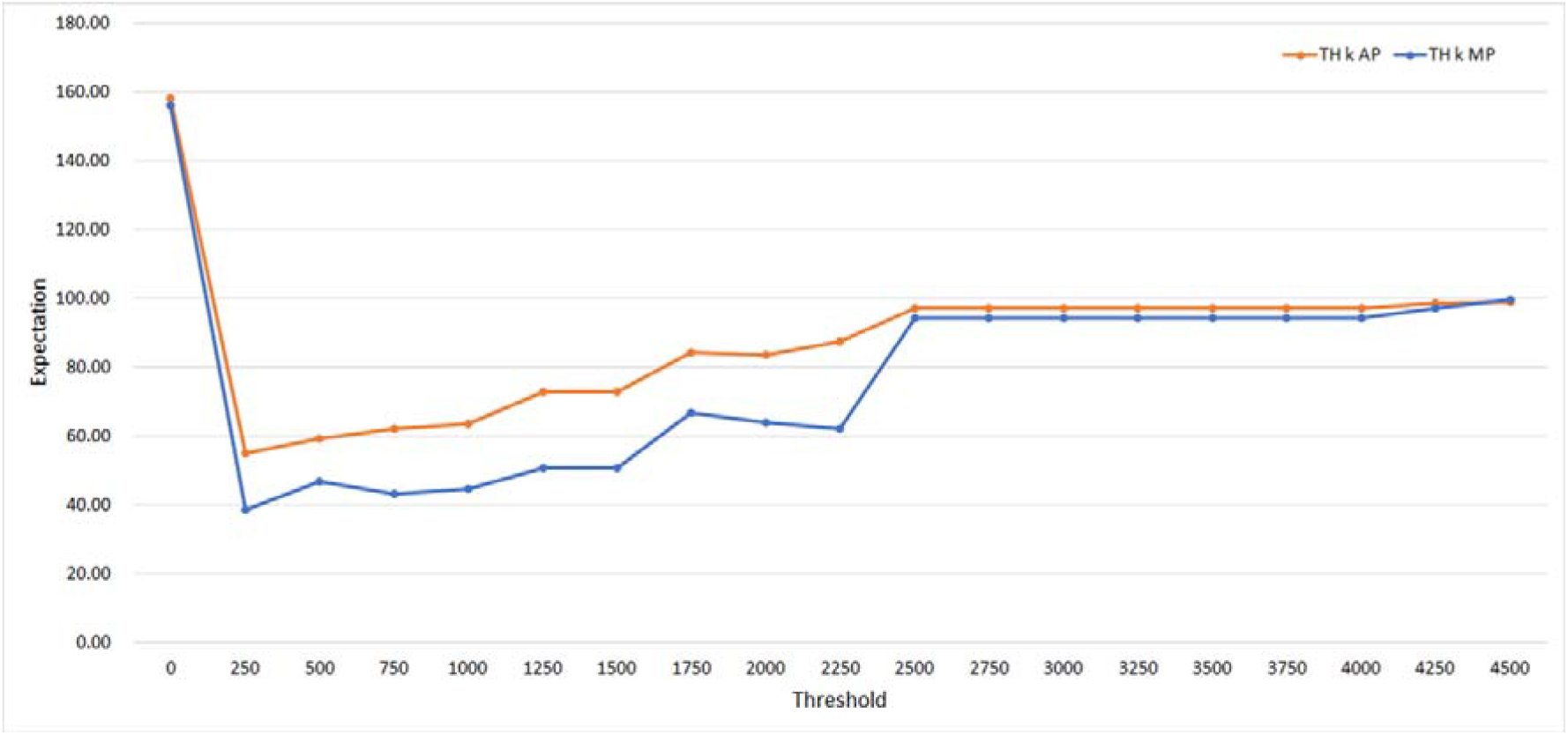
The results for TH *k* AP and TH *k* MP when the threshold was increased from 0 to 4,500 at increments of 250.

To gain a comprehensive understanding of this expectation, we conducted experiments by increasing the threshold *k* from 0 to 4,500 in increments of 250. The results are presented in Fig. 3. TH 500 AP to TH 4,500 AP consistently demonstrated the best performances in terms of coverage with a value of one, yet with relatively high median values (all results are shown in Supplementary Material 1). In contrast, OR yielded the best median but the worst coverage. TH 250 MP exhibited the highest expectation among all tested threshold settings (all results are indicated in Supplementary Material 1).

Based on this, we conducted additional experiments by incrementally increasing the threshold value around TH 250 MP from 0 to 500 in increments of 10 to determine the optimum value of *k* (Fig. 4). With *k* of 190, the expected value reached the highest performance with 38.39. However, from *k*=190 to 300, only small variations were observed, and the performance was maintained. The results of this experiment are presented in the supplementary data in Supplementary Material 2.

**Fig. 4.**
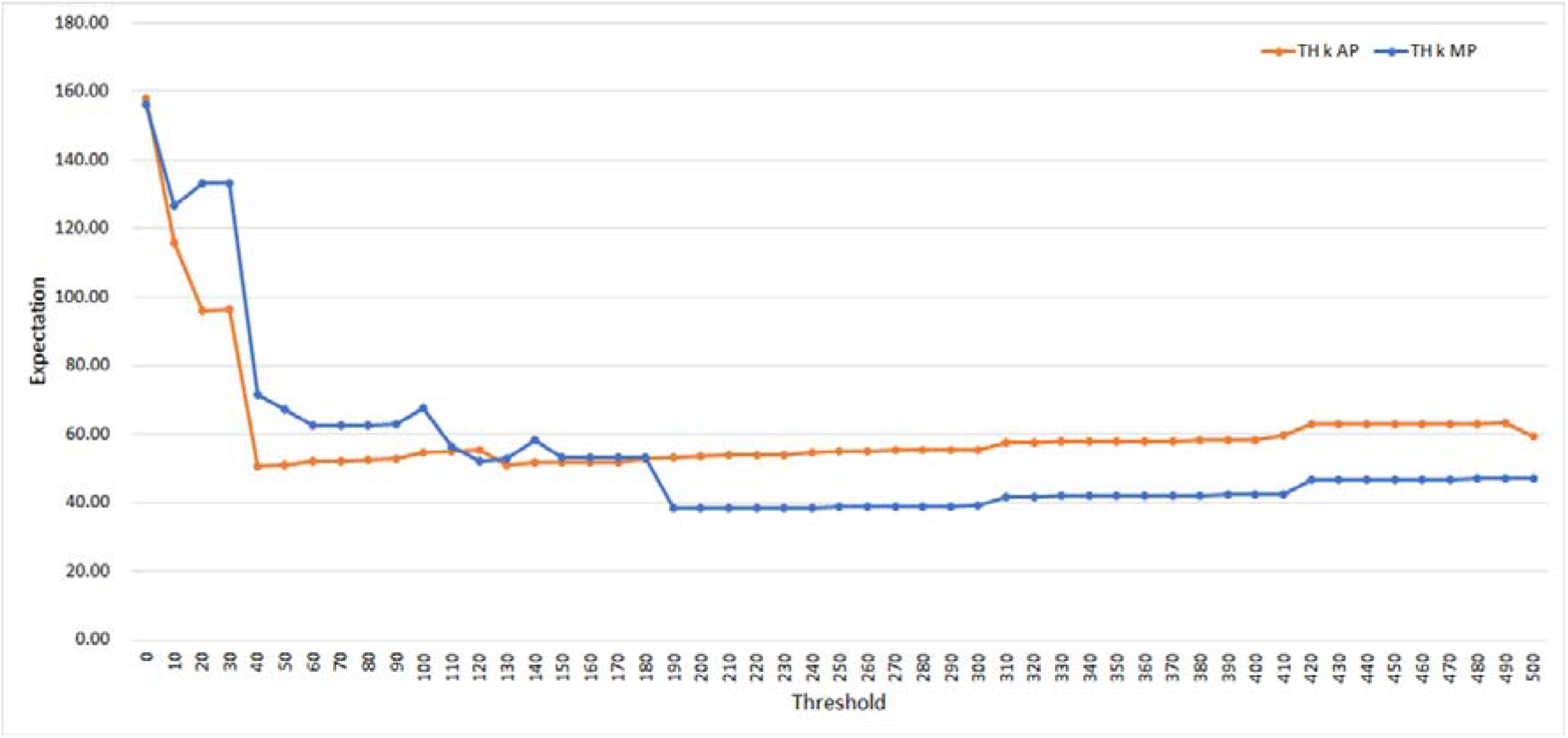
The results for TH *k* AP and TH *k* MP when the threshold was increased from 0 to 500 at increments of 10.

Table 3 shows the results for the seven expansion methods OR, 1UAP, 1UAP, 1UMP, 2UAP, and 2UMP, and the optimal thresholds TH 40 AP and TH 190 MP. Here, TH 190 MP yielded the highest expectation.

**Table 3.** The results of the experiments using OR, 1UAP, 1UMP, 2UAP, 2UMP, TH 40 AP and TH 190 MP.

|  | OR | 1UAP | 1UMP | 2UAP | 2UMP | TH 40 AP | TH 190 MP |
| --- | --- | --- | --- | --- | --- | --- | --- |
| **Coverage** | 0.4864 | 0.9324 | 0.8378 | 0.9729 | 0.8918 | 0.9595 | 0.9459 |
| **Median** | 1 | 23.5 | 10 | 60 | 18 | 31 | 14 |
| **Expectation** | 197.22 | 52.77 | 74.67 | 67.14 | 64.87 | 50.55 | 38.39 |

## Discussion

Among the seven methods, TH *k* MP exhibited the best performance, with an optimal *k* value of 190 (Supplementary Material 2). With increasing values of *k*, both the coverage of TH *k* MP and the median number of genes to be analyzed increased as well. Consequently, a trade-off between the coverage and the median exists. Users can design a VGP with their preferred coverage by selecting appropriate values of *k*.

The number of candidate genes to be analyzed may also be reduced using VGPs. If the VGP is not used, all candidate genes will need to be analyzed (median: 388). However, using VGPs designed by the TH 190 MP, the median number of candidate genes reduces to 14 if the VGPs include disease-causing genes (all results are indicated in Supplementary Material 2). Therefore, VGPs designed using the methods proposed here may be useful for gene-ranking systems such as PubCaseFinder [15].

Here, VGPs were developed using simple rolling-up and rolling-up methods with a threshold *k*. We plan to develop more sophisticated filtering methods that involve examining gene lists from surrounding and superclass disease groups, calculating similarities, and performing more refined filtering starting from the initial diagnosis stage. This approach will allow for a more tailored and precise analysis of candidate genes, considering their relevance to specific disease contexts and their functional similarities to known disease-causing genes. We will also focus on developing more sophisticated and refined filtering methods for VGPs, enabling more accurate and targeted analyses of candidate genes in the context of specific diseases.

## Conclusion

In this study, seven methods are presented to develop expanded VGPs for initial clinical diagnosis using knowledge graphs based on Mondo. The results indicated that the minimum path up with the threshold *k* method, TH *k* MP, showed the best performance in terms of expectation with a threshold of 190. This method can be used to identify candidate genes, and facilitate rare disease diagnosis efficiency. We expect that this method will be widely used by clinicians to diagnose rare diseases using NGS data.

## Data Availability

The knowledge graph data used in this study can be freely accessed via the RDF Portal https://rdfportal.org/dataset/pubcasefinder.
The data that support the findings of this study were obtained from the Hamamatsu University School of Medicine. Restrictions apply to the availability of this data, which were used under license only for the current study and thus are not publicly available. However, data are available from the authors upon reasonable request and with permission from the Hamamatsu University School of Medicine. The source code has also not yet been released, since it was developed for experimental purposes only.

## Abbreviations

1UAP: One class Up for All Paths
1UMP: One class Up for Minimum Paths
2UAP: Two classes up for all paths
2UMP: Two classes up for minimum paths
Mondo: Mondo disease ontology
NGS: Next-Generation Sequencing
OR: Original set
RDF: Resource Description Framework
TH *k* AP: All paths up with threshold *k*
TH *k* MP: Minimum path up with threshold *k*
VGP(s): Virtual Gene Panel(s)
WES: whole exome sequencing

## Supplementary Information

Supplementary Material 1

Supplementary Material 2

## Acknowledgements

We would like to thank the Semantic Web Applications and Tools for Health Care and Life Sciences (SWAT4HCLS) for providing us with space to work on this project and for valuable feedback during online sessions.

## Authors’ contributions

JS, TF, and AY: conceptualization and methodology, investigation, implementation of the algorithm, data analysis, and interpretation (statistics, evaluation, and presentation of the results). JS and TF: Data extraction. JS, TF, and HS: Data preparation. JS: Writing, original draft preparation, and system development. TF and AY: Writing – review and editing and supervision. All the authors have read and approved the final manuscript.

## Availability of data and materials

The knowledge graph data used in this study can be freely accessed via the RDF Portal https://rdfportal.org/dataset/pubcasefinder.

The data that support the findings of this study were obtained from the Hamamatsu University School of Medicine. Restrictions apply to the availability of this data, which were used under license only for the current study and thus are not publicly available. However, data are available from the authors upon reasonable request and with permission from the Hamamatsu University School of Medicine. The source code has also not yet been released, since it was developed for experimental purposes only.

## Declarations

### Ethics approval and consent to participate

Not applicable.

### Consent for publication

Not applicable.

## Authors details

^1^Database Center for Life Science, Kashiwa, Chiba, Japan

^2^Hamamatsu University School of Medicine, Hamamatsu, Japan

^3^Tokyo City University, Setagaya, Tokyo, Japan

